# Another Feature of Silicon Nanowire Field Effect Transistor Biosensor: Dynamic Detection

**DOI:** 10.1101/2020.10.04.20206532

**Authors:** Hang Chen, Lijuan Deng, Hang Li, Longchang Huang, Xiaoping Zhu, Yanfeng Jiang, Tong Wang

## Abstract

Silicon nanowire field effect transistor (SiNW-FET) biosensors are capable of label-free, real-time and biological detection with high sensitivity and specificity. However, direct observation on protein-protein interaction in blood or serum is still very difficult because of the complex physiological environment and Debye-screening effect. In order to overcome the detection obstacles, we used dialysis desalination method to purify the detection fluid and overcome Debye-screening effect. In our research, a top-down approach was proposed to fabricate the SiNW-FET, APTES-Glu chemical chain was used to link antibody to the SiNWs. And after verified the detection ability of silicon nanometer biosensors, the dynamic detection process of HbA1c was successfully realized. This could be helpful for accurate diagnosis of diabetes for clinical application. And it makes it possible to dynamic research of small biomolecules without markers.

## 1 Introduction

semiconductor nanowire field effect transistor biosensors are capable of label-free, real-time and biological detection with high sensitivity and specificity. During the past two decades, this semiconductor biosensors, either fabricated by top-down [1-2] or bottom-up methods [3], have vastly applicated ranging from medical testing, biopharmaceutical, environmental monitoring and cell research [4-8]. What’s more, due to its stable performance and simple fabrication method, silicon nanowire field effect transistor (SiNW-FET) has become the most representative semiconductor biosensorare [9-10].

Next-generation biomedical sensors will need to be sensitive to detect protein-protein interaction [11-12]. Such sensors would enable the accurate and *in-situ* detection of physiological signals. SiNW-FET could be a candidate for such kind of direct observation of protein-protein interaction because of its ultra-sensitivity and its bio-compatibility [8]. However, there are still many problems to hinder the SiNW-FET from the real application because of the Debye-screening effect [13]. The effect shields the charged proteins to be detected by the transistor. Also, it is very difficult to characterize the whole blood or serum directly because of the complex liquid with many kinds of salt ions, urea and proteins, *et al*. [14-19].

From nano-bioelectronics standpoint, The detection principle of semiconductor nano-biosensor is the electric field effect of charged biomolecules on the nano-biosensor. For example, The negatively charged protein molecules assemble around the p-type silicon nanowires, causing the internal carriers of nanowires to gather, and increasing current and conductance. The ultra-sensitivity response of SiNW-FET biosensors is related to the binding affinity, concentration and diffusion capacity of the charged biomolecules at the electrolyte solution[8, 10, 20].

Because of the Debye-screening effect, it is difficult for nanowire field effect transistor (FET) to analyse clinical samples quickly and accurately [21]. There are several methods to overcome Debye-screening effect for nanowire FET detection, including 1) using microcentrifuge filter to purify serum proteins [14] and using antigen-antibody binding reaction to purify target protein [16] to cause the removal of ions and increasing the Debye-screening distance, 2) tailoring antibodies [19] and using aptamers instead of antibodies [22-23] to shorten the distance between the target molecule and the semiconductor nanowires therefore put them within the Debye-screening distance range, 3) modifying a thin layer of biomolecule-permeable polymer on the nanowires to increase the Debye-screening distance [17], 4) High frequency measurement [18].

Although these measures can realize the detection of physiological solution by nanometer biosensor, they do not provide more value for the clinical application of semiconductor biosensors due to the complicated process and diminished the sensitivity and specificity of the device [14, 24].

Here we conduct a high-efficiency and stable biosensor for overcoming Debye-screening effect and direct analyse of the proteins in the detection solution. The dialysis desalination can weaken solution ion concentration as shown in our previous research [25]. And the Debye-screening distance is increased as the solution ion concentration decreased. In this way, the influence of the target proteins on the conductance of the device can be characterised clearly in the detection solution. What’s more, it is the first time to use nano biosensor dynamic observing the generation process of the glycosylated hemoglobin (HbA1c) and the interaction between the glycosylated hemoglobin and its antibody in the dialyzed detection solution. This will undoubtedly be the leapfrog application progress of nanosensors.

## 2 Materials and methods

The p-type silicon-on-insulator wafers (ρ:10–20 Ω cm) were purchased from Nova Electronic Materials. Polydimethylsiloxane (PDMS) was obtained from Dow Corning Co. (Michigan, USA). Phosphoric acid buffer solution (PBS), 3-aminopropyltriethoxysilane (APTES), glutaraldehyde (Glu), the CEA, IL-6 and antibodys were obtained from Sigma-Aldrich (St Louis, MO, USA). Anti-HbA1c was obtained from Wondfo (Guanzhou, China). The serum was obtained from 20 patients who were clinical suspected diabetes. All of these patients provided writtent informed consent to participate in the study. The dialysis membrane was obtained from Fresenius Medical Care (Bad Homburg, Germany).

### 2.1 Characteristic of SiNW-FET

The silicon nanowire (SiNW) transistors are fabricated using a top-down approach. All the fabrication processes and modification processes of silicon nanosensors can be found in our previous paper [25]. The biosensor is double-gate field effect tube, The schematic of the transistor structure is shown in Fig. 2a. Among them, the source and drain forms a loop through silicon nanowires. G_1_ and G_2_ represent top gate and back gate respectively. SEM image of the fabricated nanowire integrated circuit chip is shown in Fig. 2b, the width of silicon nanowires is 500 nanometers. The TEM images on the nanowire are shown in Fig. 2c, in which shows the diffraction pattern of the sample. The performance results (output curve and transfer curve) of SiNW-FET as shown in Fig.2d. This illustrate that the biosensor has good performance of electric field modulation, which is the basis of biological detection.

**Figure 1.**
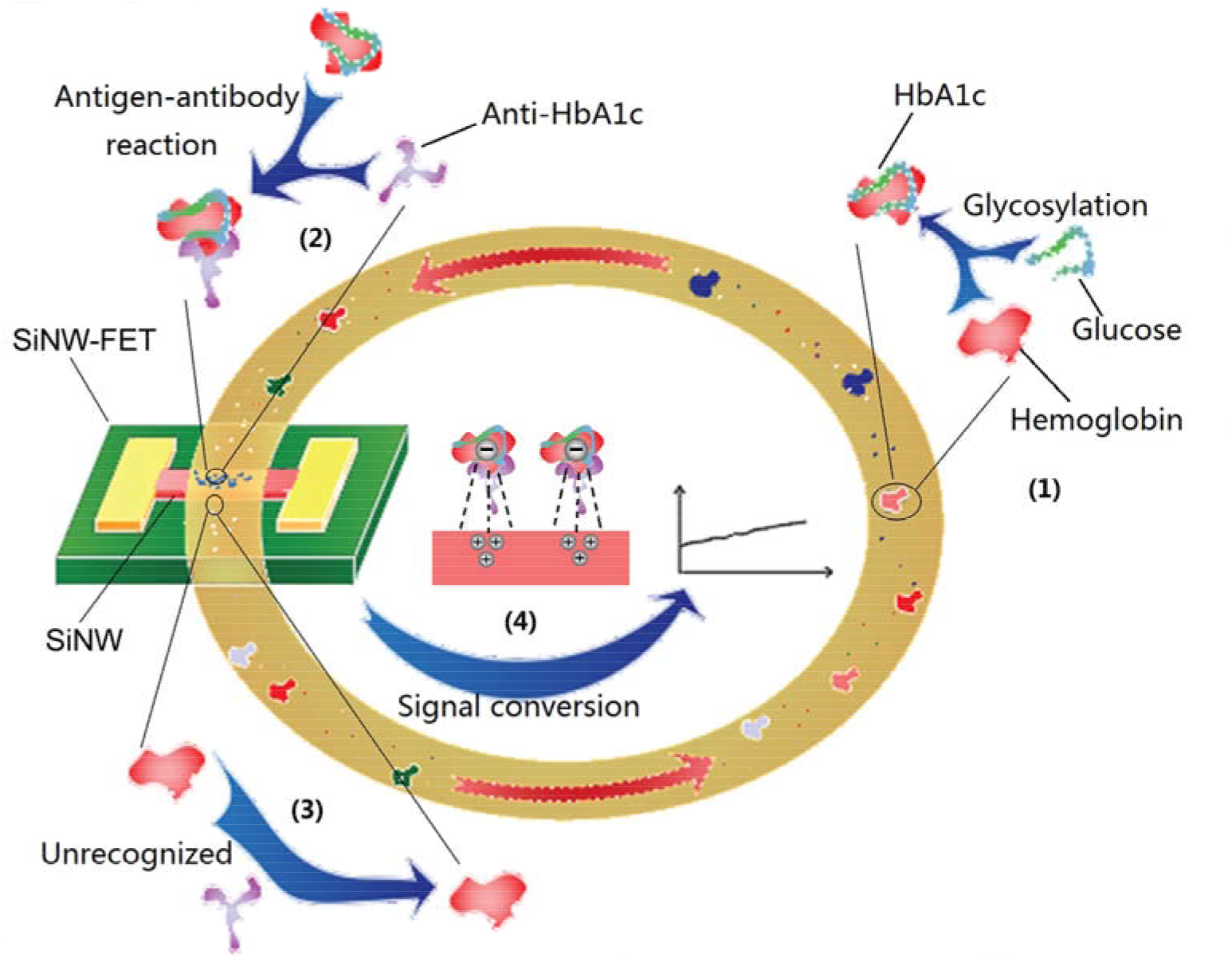
Schematic of the dynamic detection of HbA1c by Dialysis-SiNW-FET in circulating detection solution. (1) Nonenzymatic glycosylation of hemoglobin. (2) Antigen-antibody reaction of HbA1c, specific recognition and binding. (3) HbA1c antibodies could not recognize hemoglobin. (4) The negative electric field effect makes the carriers in the silicon nanowires accumulate and enhances the output current.

**Figure 2.**
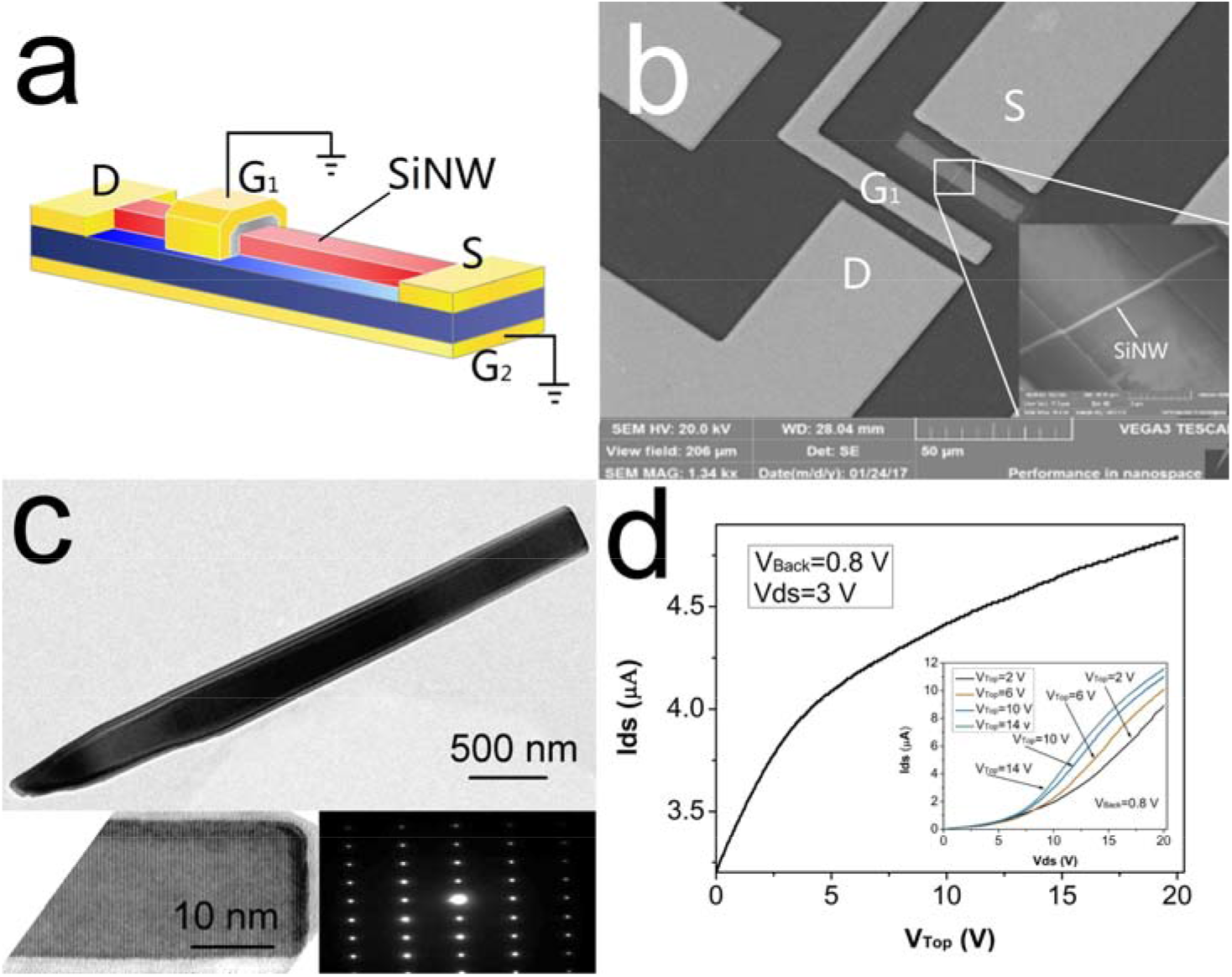
The fabricated SiNW transistor and the related characterization results. (a) The schematic of the transistor with double gate structure. (b) SEM image of the fabricated nanowire integrated circuit chip. (c) The TEM images on the nanowire. (d) The performance results (output curve and transfer curve) of SiNW-FET. Reprinted with permission from Ref [25].

### 2.2 Surface Functional Antibody Modifications

The modification of relevant antibodies on the surface of silicon nanowires is a key step to realize the specific detection of silicon nanosensor. Antibodies can specifically capture the target molecule and form a charge accumulation around the nanowires, thereby altering the output current. APTES-Glu chemical chain was used to link antibody to the SiNWs in our research. The modification process as shown in our previous paper [25]. We used the surface morphology detection technology and fluorescence development technology to prove the effectiveness of the modification method. Figure 3a-d shows the atomic force microscope figures after the modification of the CEA antibody, the morphology of nanowires and modified protein molecules can be seen in the figure. Figure 4a-d shows the picture of a device that modified the fluorescent protein. It can be seen that the fluorescent protein was successfully modified on the nanowire. These results demonstrated the good connectivity between APTES-Glu chemical chain and protein.

**Figure 3.**
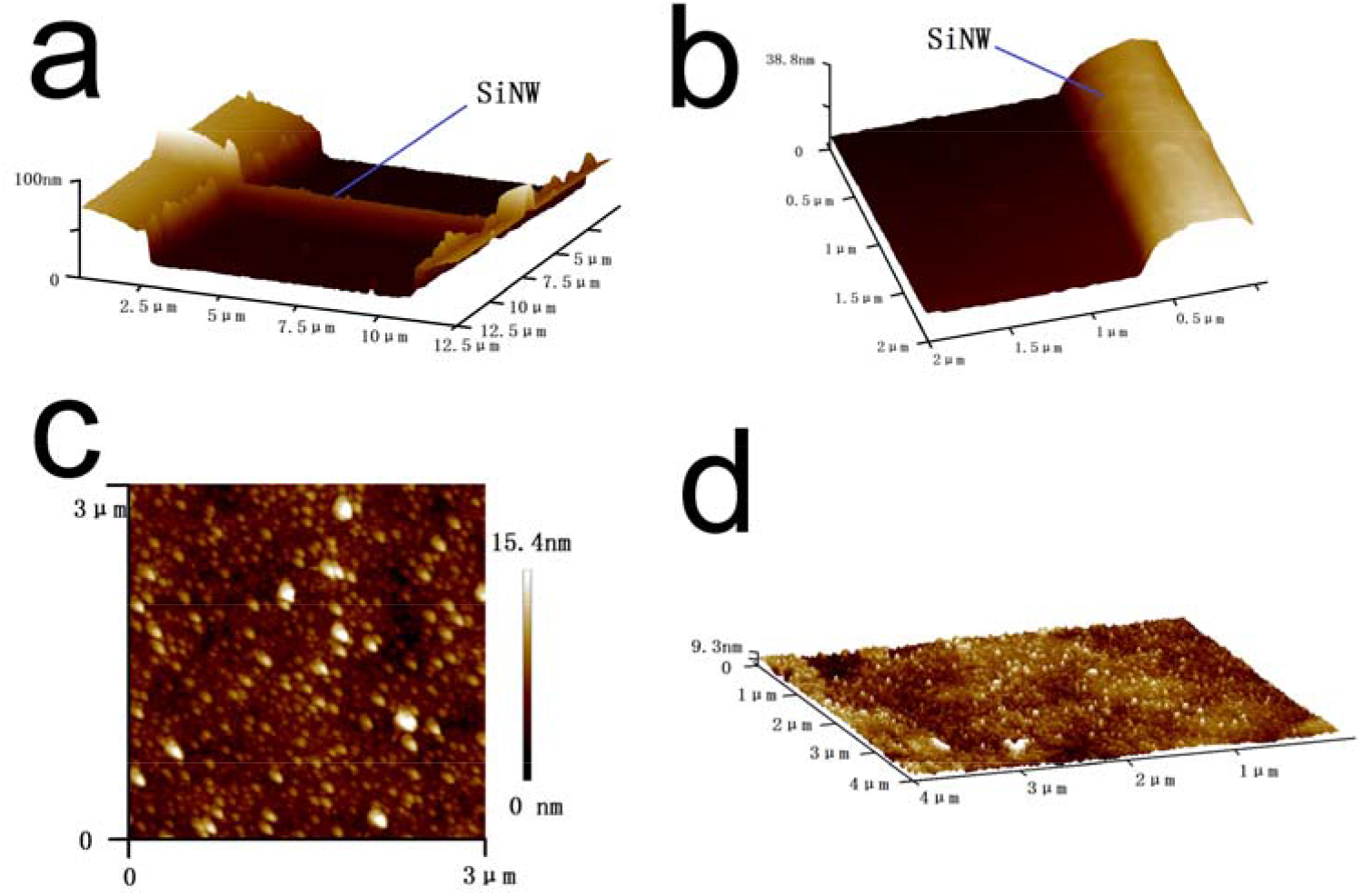
The atomic force microscope figures of SiNW after the CEA antibody modification.

**Figure 4.**
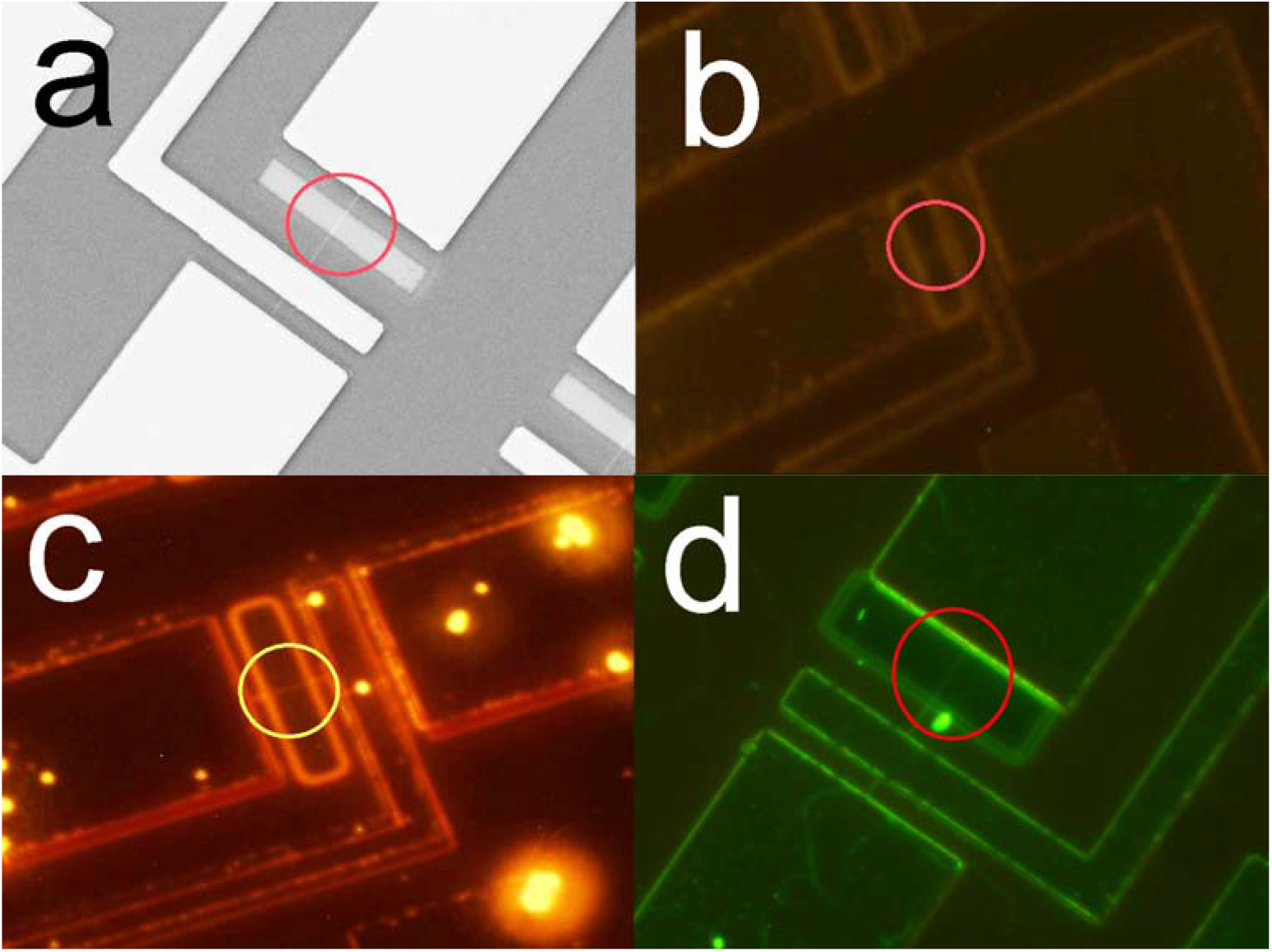
Results of APTES-Glu chemical chain modification. (a) Schematic diagram of silicon nanowire structure. (b) result of fluorescent protein solution rinsing the nanochip directly. (c d) The result of fluorescent protein solution rinsing the nanochip after modification of APTES-Glu chemical chain.

### 2.3 Overcome the Debye-screening effect

Debye-screening effect is widely existed in the detection of semiconductor nanosensor. The screening strength can be quantified by debye length (λ_D_). The stronger debye shielding effect is, the shorter λ_D_ is. And according to the Debye-screening effect principle [11, 26]:

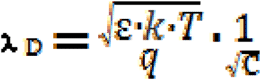

Where ε is the dielectric permittivity of the solution, k is Boltzmann’s constant, T is the temperature, q is the elementary electron charge, and c represents the ionic strength of the solution. For common physiological solution sample (i.e. blood, serum, and urine), λ_D_ is approximately 0.7 – 2.2 nm, and thus nanowire FET sensors exhibit poor sensitivity to detect large biomolecules in physiological solutions where the surface potential is completely screened at a distance of a few nanometers.

According to the formula, reducing the concentration of serum salt ions can effectively reduce the detection interference of the Debye-screening effect. The use of dialysis to remove salt ions is a simple and quick desalination process, with the benefits of high efficiency of salt ions filtration, low loss of target molecules and no change in liquid volume before and after dialysis. In this paper, a dialysis membrane with a certain aperture (10,000 D) was used to separate the detection solution from the dialyzate (deionized water). The diffusion of the salt ions is driven by the concentration difference on both sides of the membrane. A balance will be reached finally while the salt ion concentrations on both sides are the same.

Clinical hemodialysis machine is a great invention to purify the blood [27]. It can eliminate blood electrolyte disorder, remove the harmful metabolites in the blood, maintain the acid-base balance rapidly and efficiently. Therefore, hemodialysis has become an indispensable treatment in clinical treatment. The basic principle of Clinical hemodialysis machine is the filtration capacity of dialysis membrane. But the more important ability of hemodialysis machine is to control the volume of blood after dialysis. On the basis of the hemodialysis machine, we reduced the dialyzer to our ideal size. And changed the clinical equivalent osmotic fluid into deionized water, which achieved the purpose of detection solution equivalent volume desalination efficiently. Figure 5a shows the principle of dialysis desalination of the shrunken dialysis machine. Various salt ions, creatinine, uric acid and urea will be filtered by cis concentration gradient, while the protein molecules will remain in the detection solution. Figure 5b shows the relationship between the dialysis time and the concentration of salt ions in the serum. It is found that the concentration of salt ions decreased significantly by treating 1 ml serum into the machine, and continued to decline with the extension of dialysis time. After 20 s, the salt ionic concentration is about 4.5 mmol/L, reaching our ideal salt ionic concentration. And the decline trend of salt ionic concentration become slow. These results demonstrate the high efficiency of dialysis desalination. By this approach, most of the compositions in the detection solution keep unchanged while the salt ions, urea, creatinine and uric acid are removed [28][28]. More importantly, dialysis desalination breaks the limitations of traditional desalination methods, such as multiple steps, time consuming and large loss of target molecules. Therefore, dialysis desalination can obviously improve the Debye-screening distance and reduce the Debye-screening effect of SiNW-FET.

**Figure 5.**
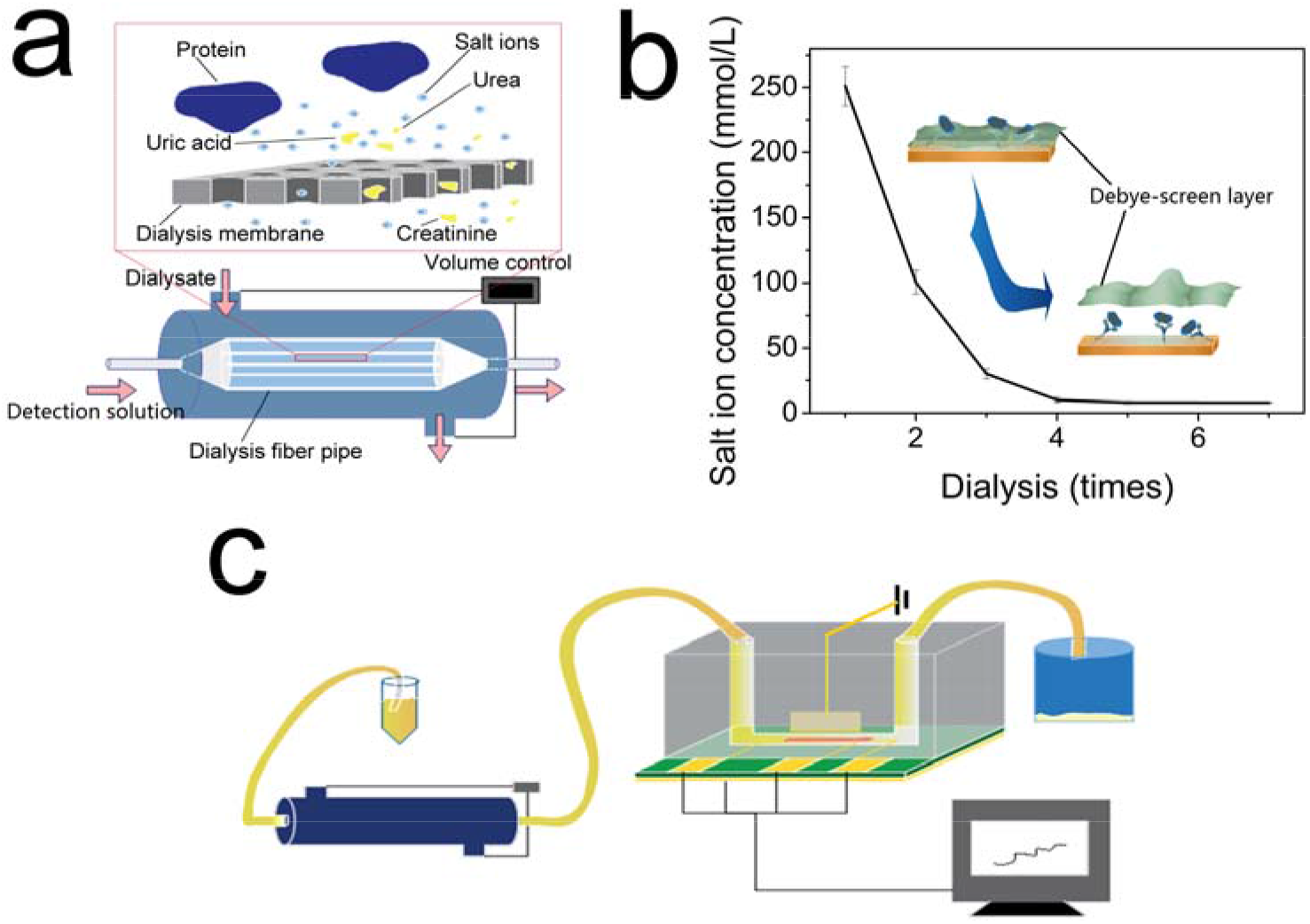
The principle of dialysis desalination to overcome Debye-screening effect. (a) Schematics of the shrunken dialysis machine. (b) Relationship between the dialysis time and the concentration of salt ions in the serum. (c) The Dialysis-SiNW-FET system.

To further achieve the real-time characterization of SiNW-FET, the Dialysis-SiNW-FET system is built-up by connecting the dialyzer with SiNW-FET in the same flowing channel, as shown in Fig. 4c. The test solution is input into the system through a silicone hose to realize real-time detection.

## 3 Results and discussion

### 3.1 Verifying the Dialysis-SiNW-FET detection ability

Phosphate buffer saline (PBS), IL-6, CEA, dog serum and human serum were used to demonstrate the specificity and sensitivity of the Dialysis-SiNW-FET. Specificity detection results of SiNW-FET as shown in Fig. 6, and the Numbers in the figure represent different solutions, which are respectively (1) PBS, (2) 5 pg/ml IL-6 standard solution, (3) 5 pg/ml CEA standard solution, (4) dog serum, (5) human serum (containing 1.25 ng/ml CEA and 5 pg/ml IL-6), (6) dog serum with 1ng/ml CEA and (7) dog serum with 50 pg/ml IL-6. CEA and IL-6 standard solution was corresponding protein dissolved in 0.02×PBS. As shown in Fig. 6a, the sensor was not modified with any antibody, and no current change was observed after injecting various protein detection solutions. The detection result after modifying CEA antibody as shown in Fig. 6b and Fig. 6c. It can be seen that only when the solution containing CEA can change the sensor current. A similar result were shown in Fig. 6d and Fig. 6e, where the sensors modified with IL-6 antibody shown a current change only when it detected solutions containing IL-6. It means that the detection specificity of the sensor depends on which antibody is modified, and the ability of relevant antibodies to recognize and capture antigens is the basis of the specific detection of SiNW-FET.

**Figure 6.**
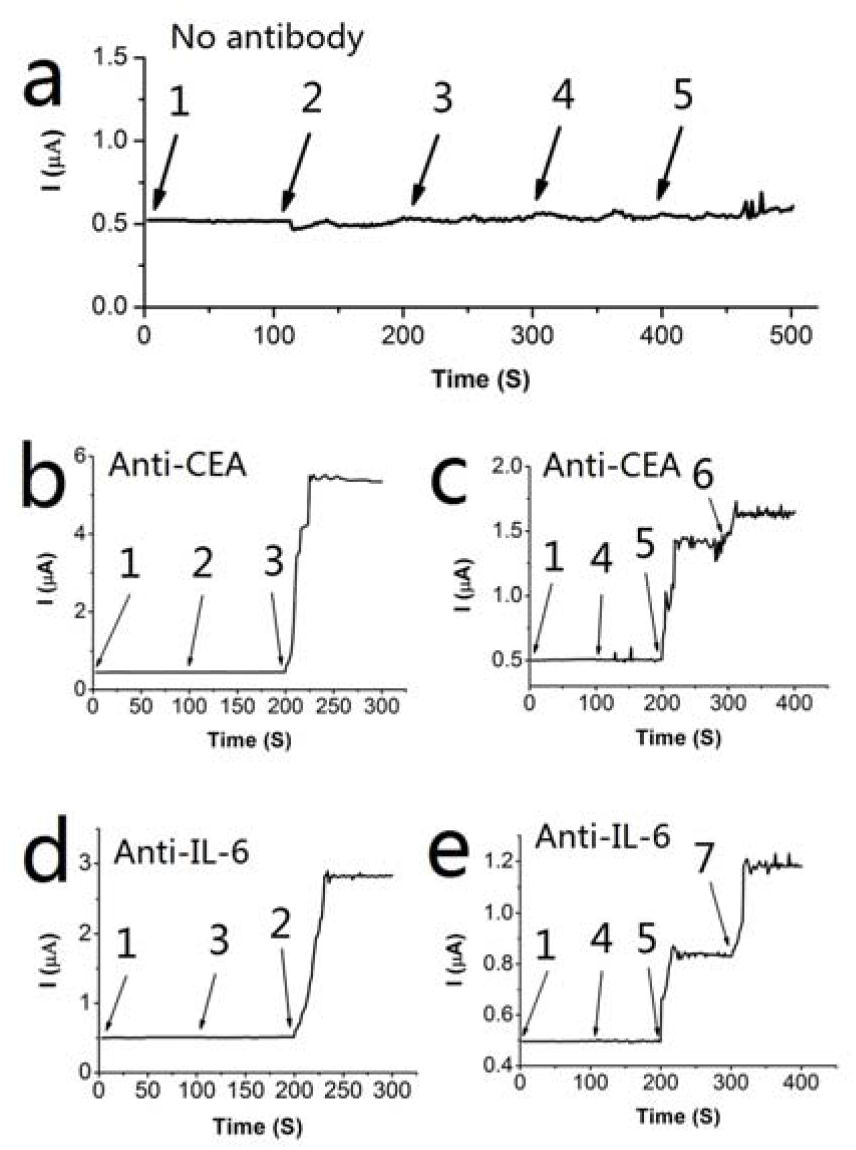
Specificity detection results of SiNW-FET. The Numbers in the figure represent different solutions, which are respectively (1) PBS, (2) 5 pg/ml IL-6 standard solution, (3) 5 pg/ml CEA standard solution, (4) dog serum, (5) human serum (containing 1.25 ng/ml CEA and 5 pg/ml IL-6), (6) dog serum with 1pg/ml CEA and (7) dog serum with 50 pg/ml IL-6. (a) When a sensor modified without antibodies, the various protein solutions did not cause a significant change in the output current value. (b c) When a sensor modified with anti-CEA, the protein solutions with CEA cause a significant change in the output current value. (d e) When a sensor modified with anti-IL-6, the protein solutions with IL-6 cause a significant change in the output current value.

Figure 7a and Figure 7b shown the sensitivity of the Dialysis-SiNW-FET. It can be seen that different concentrations of CEA standard solution lead to different output current change by the device modified with Anti-CEA. The output current increased with CEA concentration. The currents are sensitive to the concentrations of the CEA, with 3.0 μA, 5.5 μA, 7.7 μA and 8.5 μA corresponding to 10 fg/ml, 1 pg/ml, 1 ng/ml and 100 ng/ml CEA standard solutions, separately. Taking the output current change of 100 ng/ml CEA solution as I_0_, the relative percentages of the output current change at the above measured concentrations are 35.5%, 64.7% and 90.6%. Figure 7c shows the detection results of different concentrations of HbA1c. It demonstrates that the Dialysis-SiNW-FET modified with anti-HbA1c has good detection performance for HbA1c, which is the basic performance of dynamic detection of HbA1c.

**Figure 7.**
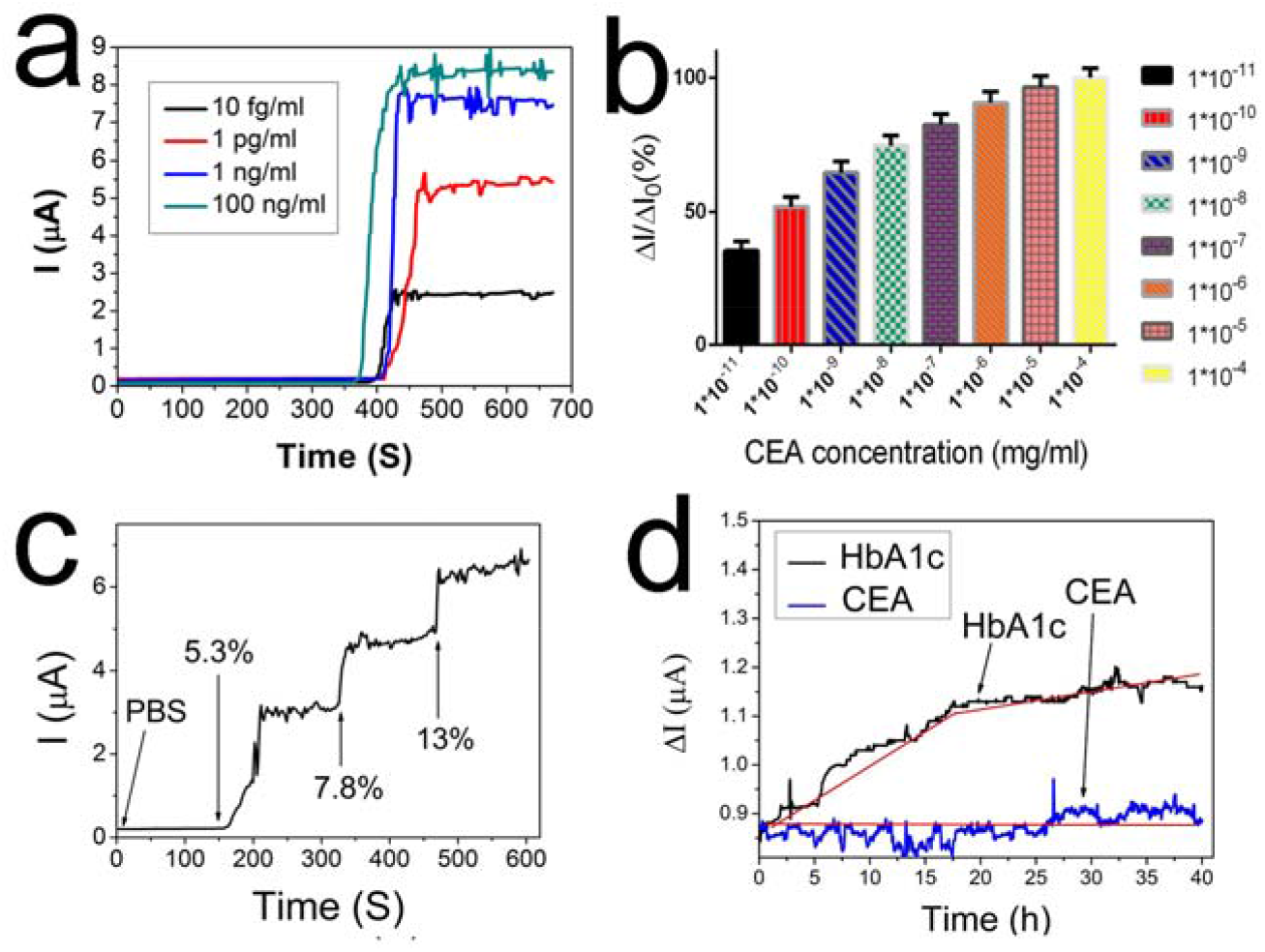
Detection results of SiNW-FET. (a) The ultra-sensitivity detection of the SiNW-FET. (b) The relative output current variation values at different CEA concentrations. (c) Detection results of HbA1c at different concentration. (d) Results of dynamic observation of HbA1c.

### 3.2 Dynamic observation of HbA1c generating

HbA1c can be used as clinic diagnosis for diabetes [29]. Recent studies show that the level of HbA1c may be more important in diabetes diagnose than the level of the blood glucose [30-32]. However, the generation of HbA1c is related to the time-consuming process of the hemoglobin glycosylation, varied from several days to several months. Therefore, the variation of the level and velocity of HbA1c over a period of time is much more meaningful than the instant value of the blood glucose level. The long term values can reflect the changing of the patient’s condition [31]. At present,the clinically determined methods of HbA1c is affinity chromatography or high performance liquid chromatography. However, these methods have limitations on detection sensitivity and complex operation process. In recent years, more novel, convenient and accurate methods have been proposed [33].

In this part, the Dialysis-SiNW-FET with prolonged Debye-screening distance is used to observe the process of the hemoglobin glycosylation. It can be considered as a new detection method with mark-free, immediate response, highly sensitive, high-specific and dynamic observation of the protein-protein interaction. The samples are taken from the clinical suspected diabetes. For the dialyzed detection solution as the precursor, its contents are purified purposely in a controlled manner [34-36]. To observe the process of the glucosylation, 20 mmol/L glucose is added into the dialyzate. The Dialysis-SiNW-FET is used to observe whether the process of the glucosylation can be conducted in the serum in real time.

For the Dialysis-SiNW-FET sensor used in the characterization, Anti-HbA1c is intentionally functionalized on the sensor’s surface. The inlet and outlet ends of SiNW are connected with a circularly sealed tube. A 40-hour continuous detection is conducted to observe the dynamic formation of HbA1c in the detection solution. Considering the possible protein degeneration in detecting serum after a long period of time, the whole experiments are conducted at 4-8 ⍰. The whole dynamic detection process is shown in Fig. 1. When the detection solution is detected for the first time, the functionalized anti-HbA1c can capture the HbA1c in it, so that the output current changed immediately and remains constant at a certain level. At this point, hemoglobin (Hb) is not recognized by anti-HbA1c. But Hb can react with glucose to form HbA1c slowly. The slowly generated HbA1c increases the HbA1c concentration in the circulating detection solution, thus increasing the output current slowly. The corresponding results are shown in Fig. 5d. it can be seen that the increment of the current corresponds to the interaction between the new generated HbA1c and the anti-HbA1c. This is the first time that the SiNW-FET is used for the real-time dynamic monitoring of continuously changing substances. In the short term, monitoring the speed and amount of HbA1c changes will undoubtedly play a good guiding role in judging the changes of diabetes patients’ conditions. It is also helpful to guide clinicians to take appropriate treatment at the right time.

## 4 Conclusion

Biosensors are capable of label-free, real-time chemical and biological detection with high sensitivity and spatial resolution in biomolecular detection. As the representative of semiconductor biosensors, silicon nano-biosensors have been proved to have good biomolecular detection ability. Although the Debye-screening effect hindered the clinical detection application of semiconductor biosensors, many research groups have eliminated the interference of the Debye-screening effect by various means. In the previous study, our research group removed the salt ions in the serum by dialysis desalination method, overcame the Debye-screening effect, and successfully detected the tumor markers in the serum by the SiNW-FET. What’s more, we found that the another ability of semiconductor biosensors: dynamic detection. which is different from other biomolecular detection technology. We used the slowly generated HbA1c in blood as the target biomolecule for the dynamic detection research of the silicon nanometer biosensor, and proved the dynamic detection ability of the SiNW-FET. This dynamic detection capability has significant clinical application potential. It is not only related to the research of the disease development process of diabetes, but also applicable to the future monitoring of blood indicators of seriously ill patients. This research confirms the detection capability of the SiNW-FET and broadens the application field of biosensors.

## Data Availability

all data in the manuscript is available

## Acknowledgments

National Science Foundation (81702592), National Sciences Foundation of Jiangsu Province (BK20160198). Major projects of Wuxi Health Committee.

